# Comparison of early ocular biological parameters in preterm infants with or without Retinopathy of Prematurity

**DOI:** 10.64898/2026.05.14.26353221

**Authors:** Panpan Ma, Qiong Wu, Wei Xin, Le Zhang

## Abstract

**Purpose:** Comparison of ocular parameters (ACD, AL, LT, VL, CCT, ASD, LC, LT/ACD) in preterm infants with retinopathy after treatment, those with spontaneous regression, and those without retinopathy, at `postmenstrual` (ages of 0 (40 weeks), 3, and 6 months.

**Methods:** Cross-sectional study. This research involved 297 premature infants assigned to three groups based on fundus results and intravitreal injection therapy: an ROP post-injection group, an ROP spontaneous regression group, and a non-ROP group. Axial length (AL), anterior chamber depth (ACD), lens thickness (LT), and vitreous length (VL) were assessed in all three groups using a corneal thickness meter at post menstrual ages (PMA) of 0, 3, and 6 months. Derived parameters—ASD (( ACD + LT), LC (( ACD + LT/2), and LT/ACD—were subsequently calculated. A one-way ANOVA analysis revealed statistically significant differences in these ocular parameters among the groups (P < 0.05).

**Results:** Significant differences emerged in anterior chamber depth (ACD) and lens thickness (LT) between the ROP post-injection group, ROP spontaneous regression group, and non-ROP group at 0, 3, and 6 (months postmenstrual age (PMA). At 0 months PMA: ACD (F=4.33, P=0.014), LT (F=5.45, P=0.005). At 3 months PMA: ACD (F=17.20, P<0.01), LT ( F=15.23, P<0.01). At 6 months PMA: ACD ( F=17.89, P<0.01), LT ( F=17.21, P<0.01). Central corneal thickness (CCT) also differed significantly among the three groups at 0 months PMA (P<0.01). All ocular parameters correlated significantly with Postmenstrual Age, with CCT and LT showing a negative correlation. Before 6 months PMA, axial length (AL) and vitreous length (VL) increased significantly, and ACD deepened significantly across all groups (P<0.05). However, LT exhibited no significant change within the ROP group (post-injection group P=0.4; spontaneous regression group P=0.33). No significant differences existed in any ocular parameters between the ROP post-injection group and the ROP spontaneous regression group (P>0.05).

**Conclusions:** Before 6 months of postmenstrual age (PMA), axial length (AL), vitreous length (VL), and anterior chamber depth (ACD) were increased between the ROP group and non-ROP group; lens thickness (LT) remained unchanged in the ROP group but increased in the non-ROP group. The injection group and the spontaneous regression group showed no significant differences. The primary factors influencing anterior segment development were birth weight (BW), gestational age (GA), and postmenstrual age (PMA).

## Introduction

With the improvement of neonatal intensive care technology, more premature infants with younger gestational age and lower body weight survive, resulting in an increase in the prevalence of ROP. Retinopathy of prematurity (ROP) is a kind of retinal vascular disease of premature infants caused by incomplete vascularization and abnormal `fibrovascular proliferation`[1-3]. Both prematurity and ROP can disrupt the normal development of ocular structures, including the retina, anterior segment, and optic nerve, potentially leading to long-term visual impairment and an increased risk of amblyopia. Visual function mainly depends on the matching of eye biological parameters, Many researchershave found that the development of myopia in premature infants may be related to ocular parameters, such as axial length and anterior chamber depth [4-6]. Others tended to believe its association with the corneal curvature and refractive power of the lens [7-9]. Current studies focused more on premature infants with ROP or whitout ROP. Our study increased the ROP spontaneous regression group and identified the presence or absence of treatment and ROP effects on ocular parameters when early ocular parameter changes occurred, In the present study, we investigated the ocular parameters in preterm infants with retinopathy after treatment and in preterm infants with spontaneous regression and without preterm retinopathy at `Postmenstrual Age` ( PMA) of 0 month( 40 weeks), 3 month and 6 month. To compare and analyze the ocular biological parameters in preterm infants with retinopathy of prematurity (ROP).

### 1.1 General information

A total of 297 premature infants who received routine ROP screening in the Department of Ophthalmology, Northwest Women and Children’s Hospital were selected from 2018/01/01-2023/01/31. Inclusion criteria: gestational age at birth (GA) was less than 37 weeks. There were no other eye diseases (such as congenital glaucoma, congenital cataract, keratopathy or other fundus diseases, etc.), craniofacial deformities and history of brain injury. Exclusion criteria: patients with stage 4 and stage 5 of ROP needed re-injection of razumab(Lucentis Novartis) and / or laser during follow-up.

This study followed the Helsinki Declaration and was approved by the Ethics Committee of Northwest Women and Children’s Hospital (the branch of the Ethics Committee of the Medical Department of Xi’an Jiaotong University). All the caregivers signed informed consent.

### 1.2 Method

1.2. 1

There are three groups

GroupA. The eyes that underwent rzumab injection (98 Diagnoses: Type 1 ROP 68, A-ROP 30; Or zone I Type 1 ROP 30 and zon2 Type 1 ROP 38 )

GroupB. Eyes with spontaneous regression of ROP(88)

GroupC. Eyes without ROP(113)

1.2.2

All infants underwent portable slit-lamp examination, RetCam3 (Clarity Medical Systems, California, USA), and A-scan ultrasound biometry (Topcon, Tokyo, Japan). Their growth parameters,including GA, BW and PMA were also recorded at each examination. The screening examination for ROP followed the guidelines recommended by the Royal College of Paediatrics and Child Health and the Royal College of Ophthalmologists, United Kingdom.15 Ultrasonography is widely used for clinical examinations of infants because it is safe and non-invasive.Ultrasonic AL measurements have been used as an indicator of eye development.The ocular biometric parameters (AL, ACD, LT and VL) were measured using A-scan ultrasound biometry. In our study, some of the participants were measured multiple times at different GA (e.g. the same subject was measuredat 32 weeks and again 2 weeks later), while other participants were only measured once.The ocular biometric parameters (AL, ACD, LT and VL) were measured using A-scan ultrasound biometry. In our study, some of the participants were measured multiple times at different GA ( e. g. the same subject was measured at 40 weeks and again 52 weeks later), while other participants were only measured once.One drop of Proparacaine Hydrochloride Eye Drops was instilled in each eye 1 to 2 min before ultrasonography. The probe was placed lightly on the center of the cornea perpendicular to its axis. The average value of at least 10 measurements for AL, ACD, LT and VL were recorded for each eye and were expressed in millimeters. All examinations were performed by the same experienced examiner. After that, 0.5% tropicamide with 0.5% phenylephrine were used to dilate the pupils. One drop was instilled in each eye every 10 minutes for a total of4 to 6 doses, 1 hour before the ROP examination.

### 1.2.3 Eye Development Patterns in Normal Children

full-term infants, most of the growth and development of the eye occurs in the first year of life; the growth curves of the (intra-)ocular dimensions AL, Dc, ACD int and VCD appear almost identical up to 1 year of age,A majority of axial length elongation takes place in the frst 3 to 6 months of life and a gradual reductionof growth over the next two years, The total anterior chamber depth (ACDtot) increases from 2.46 mm at birth to 3.59 mm at 20 years of age. LT sees three growth phases: initially, it undergoes a rapid increase like any other ocular structure until it is about 3.80 mm at birth, While unexpected,such an early increase has been reported previously for in vitro bovine lenses, followed by a decrease to 3.50 mm at 10– 14 years and finally a continuous increase during the teenage and adult years. [11-12].

1.2.3

ROP diagnostic criteria: according to the ICROP-3 criteria `ROP` study and the diagnostic criteria of the 2021 International Classification of Retinal Diseases of prematurity [13]: ( 1 ) Type 1 pre-threshold ROP (also known as high risk threshold ROP): region I, any ROP Plus disease, Plus, that is, posterior pole retinal vasodilation, tortuosity involving at least 2 quadrants; region I, stage 3 ROP without Plus disease Zone II, stage 2 or stage 3 ROP with Plus lesions; `Type 1 prethreshold ROP`: without Plus lesions occurred in area I, including stage 1 or stage 2 ROP; If they occurred in area II, they were stage 3 ROP. (2) threshold ROP: there was Plus lesion, proliferation of extraretinal fibrovascular vessels in area I or II for 5 hours or more than 8 hours.

### 1.2.5 Treatment method

Children with pre-high risk threshold and threshold ROP need to be treated within 72 hours. All the children with ROP were treated with intravitreal injection of razumab (Ranibizumab) in the operating room. `injections were performed under sterile conditions in the operating room`. 0.2mg (0.02mL) razumab was injected into the vitreous cavity. In this study, both eyes of children with ROP were treated with intravitreal injection of razumab. In this study group, all vitreous injections were performed by the same senior fundus physician.

### 1.2.6 Follow-up observation

3 days, 1 week and 2 weeks after vitreous injection, the interval of reexamination was determined according to the fundus condition, usually 1-3 weeks, at least 6 months. During the follow-up, the complications such as infection and bleeding and the prognosis of postoperative fundus lesions were observed.

#### 1.3 Statistical analysis

Statistical analysis was performed using SPSS statistical software version 23.0 (IBM Corp, Armonk, NY, USA). The values obtained for all parameters were expressed as mean ± SD. After normality testing (Shapiro-Wilk),The t-test was performed on the data that were consistent with the normal distribution The t-test was used to determine whether the differences between right or left eye and male or female. The data of the right eye of each subject were chosen for analysis to avoid methodology bias from using both eyes.One-way analysis of variance (ANOVA) and post hoc pair-wise comparison were used to analyze the differences in the parameters in the groups (ACD、LT、VL、AL 及 CCT、ASD (ACD+LT), LC (ACD+LT/2) and LT/ACD were calculated), post-hoc tests followed ANOVA.

## 2. Result

### 2.1 Basic information

A total of 297 premature infants were included in the study, including 156 males and 141 females. There were 184 preterm infants diagnosed with ROP and 113 preterm infants without ROP. The average birth weight and gestational age were 1139.38±224.57g and 28.46±1.73 weeks in group, 1628.47±469.87g and 31.82 ±2.52 weeks in group B, and 1836.04 ±437.56g and 33.09±2.06 weeks in group C. Among the premature infants with ROP, there were 27 cases of zone I `stage` 2-3, 67 cases of zone II `stage` 3, 12 cases of zone II `stage` 2, 5 cases of zone II `stage` 1, and 70 cases of zone III `stage` 1.

**Table 1.**
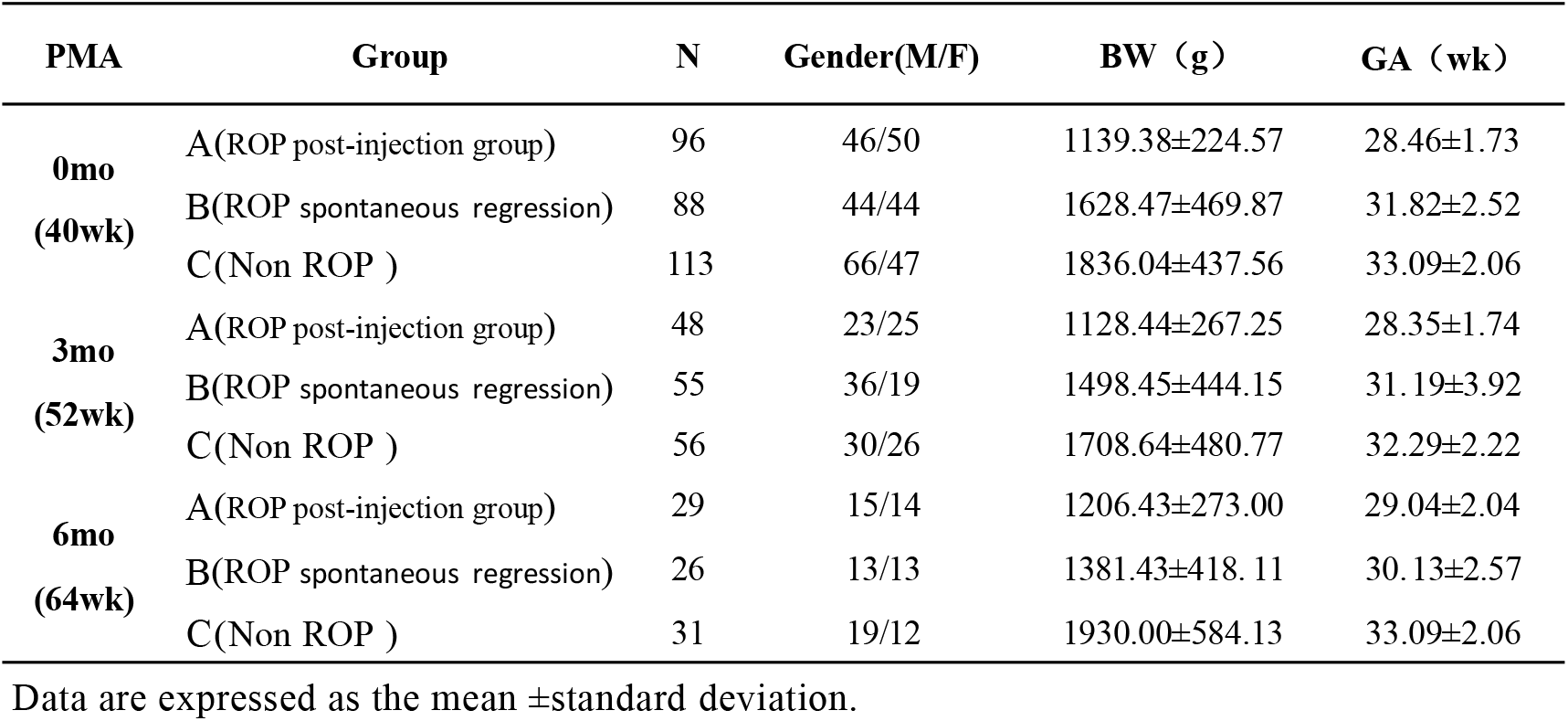
Demographic data of the study group

| PMA | Group | N | Gender(M/F) | BW (g) | GA (wk) |
| --- | --- | --- | --- | --- | --- |
| 0mo<br>(40wk) | A(ROP post-injection group) | 96 | 46/50 | 1139.38±224.57 | 28.46±1.73 |
| | B(ROP spontaneous regression) | 88 | 44/44 | 1628.47±469.87 | 31.82±2.52 |
| | C(Non ROP ) | 113 | 66/47 | 1836.04±437.56 | 33.09±2.06 |
| 3mo<br>(52wk) | A(ROP post-injection group) | 48 | 23/25 | 1128.44±267.25 | 28.35±1.74 |
| | B(ROP spontaneous regression) | 55 | 36/19 | 1498.45±444.15 | 31.19±3.92 |
| | C(Non ROP ) | 56 | 30/26 | 1708.64±480.77 | 32.29±2.22 |
| 6mo<br>(64wk) | A(ROP post-injection group) | 29 | 15/14 | 1206.43±273.00 | 29.04±2.04 |
| | B(ROP spontaneous regression) | 26 | 13/13 | 1381.43±418.11 | 30.13±2.57 |
| | C(Non ROP ) | 31 | 19/12 | 1930.00±584.13 | 33.09±2.06 |

Data are expressed as the mean ± standard deviation.

### 2.2 Eye biometric parameters

The eye parameters were compared in PMA 0 month (PMA40 week). Group A vs group B was set to P1, while group B vs group C was set to P2, and group A vs group C was set to P3 . In P1, only LC had significant difference( P= 0 . 03 ). ACD, LT, CCT, ASD and LC had significant difference in P2 and P3(ACD:P2=0.009,P3=0.02; LT:P2=0.005,P3=0.006;CCT:P2<0.01,P3<0.01;ASD:P2< 0.01,P3<0.01;LC:P2=0.028,P3 =0.029). (Table 2)

**Table 2.**
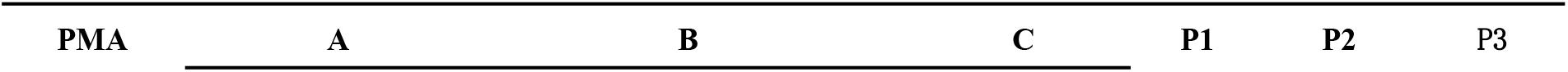
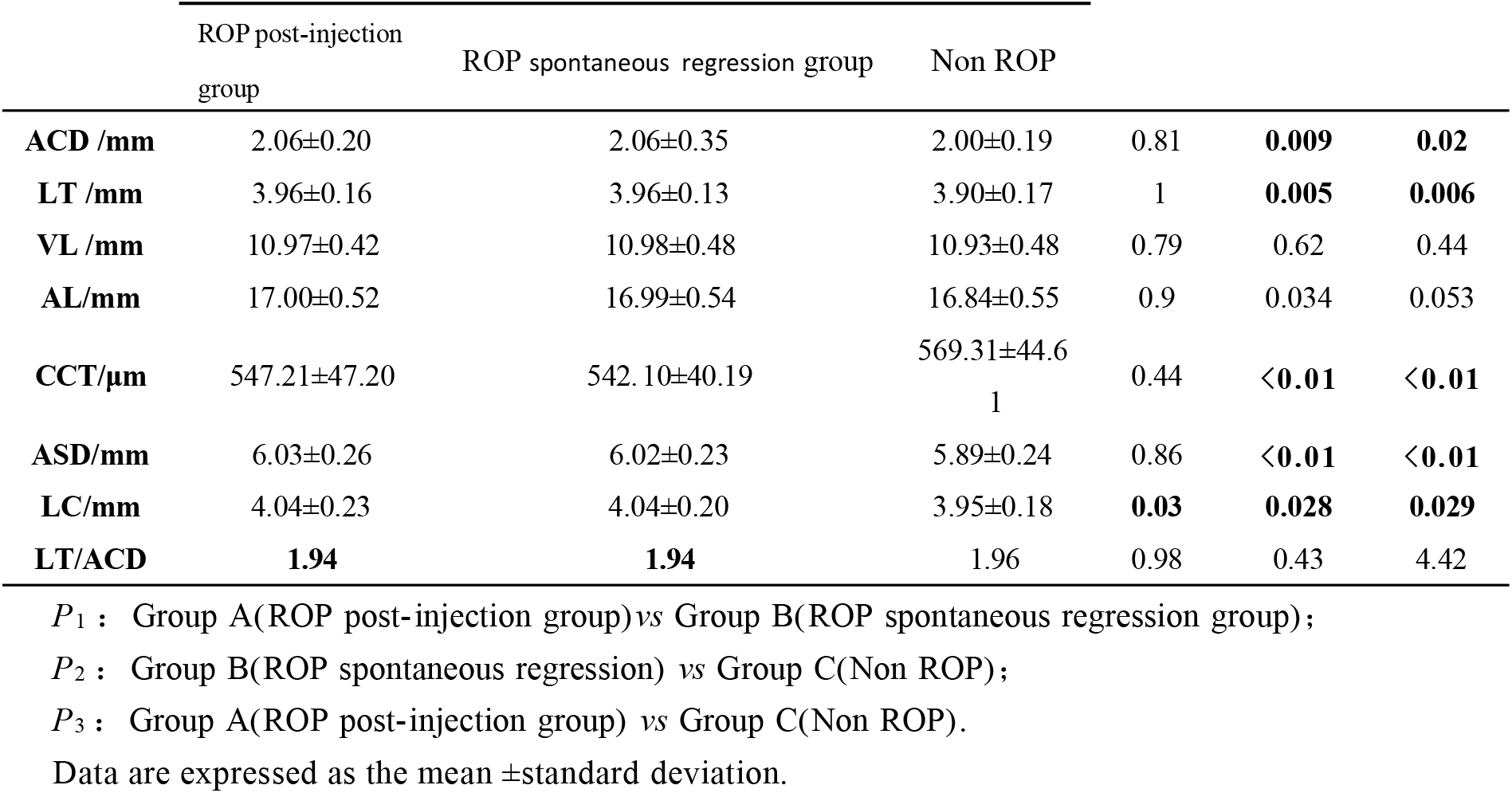
Comparison of ocular biological parameters among three groups of preterm infants with PMA in 0 month

|  | ROP post-injection<br>group | ROP spontaneous regression group | Non ROP |  |  |  |
| --- | --- | --- | --- | --- | --- | --- |
| **ACD /mm** | 2.06±0.20 | 2.06±0.35 | 2.00±0.19 | 0.81 | **0.009** | **0.02** |
| **LT /mm** | 3.96±0.16 | 3.96±0.13 | 3.90±0.17 | 1 | **0.005** | **0.006** |
| **VL /mm** | 10.97±0.42 | 10.98±0.48 | 10.93±0.48 | 0.79 | 0.62 | 0.44 |
| **AL/mm** | 17.00±0.52 | 16.99±0.54 | 16.84±0.55 | 0.9 | 0.034 | 0.053 |
| **CCT/μm** | 547.21±47.20 | 542.10±40.19 | 569.31±44.6<br>1 | 0.44 | **<0.01** | **<0.01** |
| **ASD/mm** | 6.03±0.26 | 6.02±0.23 | 5.89±0.24 | 0.86 | **<0.01** | **<0.01** |
| **LC/mm** | 4.04±0.23 | 4.04±0.20 | 3.95±0.18 | **0.03** | **0.028** | **0.029** |
| **LT/ACD** | **1.94** | **1.94** | 1.96 | 0.98 | 0.43 | 4.42 |

$P_1$ : Group A(ROP post-injection group) vs Group B(ROP spontaneous regression group);
$P_2$ : Group B(ROP spontaneous regression) vs Group C(Non ROP);
$P_3$ : Group A(ROP post-injection group) vs Group C(Non ROP).
Data are expressed as the mean ±standard deviation.

The eye parameters were compared in PMA 3 month.There was no significant difference in eye parameters in P1 (P>0.05). There were significant differences in ASD,LT and LT/ASD in P2 and P3(ACD:P2<0.01,P3<0.01;LT:P2<0.01,P3<0.01;LT/ACD:P2<0.01,P3<0.01).There was no significant difference in LC in P2(P=0.87), but there was significant difference in LC in P3(P=0.001).(Table 3)

**Table 3.** Comparison of ocular biological parameters among three groups of preterm infants with PMA in 3 month

| PMA | A | B | C | P1 | P2 | P3 |
| --- | --- | --- | --- | --- | --- | --- |
|  | ROP post-injection group | ROP spontaneous regression group | Non ROP |  |  |  |
| **ACD /mm** | 2.49±0.22 | 2.45±0.25 | 2.22±0.29 | 0.55 | **<0.01** | **<0.01** |
| **LT /mm** | 3.94±0.13 | 3.99±0.20 | 4.14±0.17 | 0.23 | **<0.01** | **<0.01** |
| **VL /mm** | 12.03±0.67 | 12.17±0.56 | 12.25±0.53 | 0.24 | 0.06 | 0.46 |
| **AL/mm** | 18.46±0.74 | 18.63±0.65 | 18.60±0.63 | 0.21 | 0.33 | 0.76 |
| **CCT/μm** | 532.85±41.98 | 523.80±38.41 | 519.50±41.53 | 0.26 | 0.1 | 0.58 |
| **ASD/mm** | 6.43±0.24 | 6.45±0.27 | 6.37±0.30 | 0.78 | 0.17 | 0.09 |
| **LC/mm** | 4.47±0.22 | 4.46±0.24 | 4.30±0.27 | 0.29 | 0.87 | **0.001** |
| **LT/ACD** | 1.6 | 1.64 | 1.89 | 0.35 | **<0.01** | **<0.01** |

Data are expressed as the mean ±standard deviation.

The eye parameters were compared in PMA 6 month.There was no significant difference in all eye parameters in P1. ACD,LT,LC and LT/ACD had significant differences in P2 and P3 (ACD:P2<0.01,P3<0.01; LT:P2=0.013,P3 <0.01;LC:P2= 0.009,P3 =0.001;LT/ACD:P2<0.01,P3 < 0.01).(Table 4)

**Table 4.** Comparison of ocular biological parameters among three groups of preterm infants with PMA in 6 month

| PMA | A | B | C | P1 | P2 | P3 |
| --- | --- | --- | --- | --- | --- | --- |
|  | ROP post-injection group | ROP spontaneous regression group | Non ROP |  |  |  |
| ACD /mm | 2.68±0.20 | 2.59±0.35 | 2.30±0.19 | 0.20 | <0.01 | <0.01 |
| LT /mm | 3.91±0.18 | 4.02±0.34 | 4.29±0.23 | 0.13 | 0.013 | <0.01 |
| VL /mm | 12.60±0.65 | 12.80±0.81 | 12.94±0.58 | 0.27 | 0.41 | 0.05 |
| AL/mm | 19.20±0.70 | 19.39±0.88 | 19.49±0.67 | 0.32 | 0.62 | 0.13 |
| CCT/μm | 521.61±41.75 | 515.61±32.26 | 522.35±26.35 | 0.51 | 0.45 | 0.93 |
| ASD/mm | 6.59±0.21 | 6.61±0.29 | 6.59±0.25 | 0.81 | 0.75 | 0.94 |
| LC/mm | 4.64±0.19 | 4.60±0.28 | 4.45±0.20 | 0.53 | 0.009 | 0.001 |
| LT/ACD | 1.47 | 1.58 | 1.88 | 0.07 | <0.01 | <0.01 |

Data are expressed as the mean ±standard deviation.

There were statistically significant differences in ACD,VL,AL,CCT,ASD and LC in three agroups at PMA 0,3,6 month(ACD,VL,AL,ASD and LC in three groups, P<0.01; CCT: Group A :P= 0.017; group B :P=0.001; group C :P <0.01).There were significant differences in LT in group C (P < 0 . 01 ) . There was significant difference in LT/ ACD between group A and group B ( group A: P < 0.01; group B: P < 0.01).The LT of premature infants with ROP (group A and B) had no development, and the LT of premature infants without ROP (group C) gradually thickened at the post modification age of 0-6 months.(Table 5)

**Table 5.** Comparison of eye parameters in three age groups of three groups premature infants

|  | Group | 0mo | 3mo | 6mo | F | P |
| --- | --- | --- | --- | --- | --- | --- |
| ACD /mm | A (ROP post-injection group) | 2.06±0.20 | 2.49±0.22 | 2.68±0.20 | 114.42 | <0.01 |
|  | B (ROP spontaneous regression group) | 2.06±0.35 | 2.45±0.25 | 2.59±0.35 | 74.36 | <0.01 |
|  | C (Non ROP ) | 2.00±0.19 | 2.22±0.29 | 2.30±0.19 | 40.73 | <0.01 |
| LT /mm | A (ROP post-injection group) | 3.96±0.16 | 3.94±0.13 | 3.91±0.18 | 0.91 | 0.4 |
|  | B (ROP spontaneous regression group) | 3.96±0.13 | 3.99±0.20 | 4.02±0.34 | 1.11 | 0.33 |
|  | C (Non ROP ) | 3.90±0.17 | 4.14±0.17 | 4.29±0.23 | 63.23 | <0.01 |
| VL /mm | A (ROP post-injection group) | 10.97±0.42 | 12.03±0.67 | 12.60±0.65 | 126.68 | <0.01 |
|  | B (ROP spontaneous regression group) | 10.98±0.48 | 12.17±0.56 | 12.80±0.81 | 139.3 | <0.01 |
|  | C (Non ROP ) | 10.93±0.48 | 12.25±0.53 | 12.94±0.58 | 248.85 | <0.01 |
| AL/mm | A (ROP post-injection group) | 17.00±0.52 | 18.46±0.74 | 19.20±0.70 | 178.15 | <0.01 |
|  | B (ROP spontaneous regression group) | 16.99±0.54 | 18.63±0.65 | 19.39±0.88 | 201.79 | <0.01 |
|  | C (Non ROP ) | 16.84±0.55 | 18.60±0.63 | 19.49±0.67 | 34.1 | <0.01 |
| CCT/μm | A (ROP post-injection group) | 547.21±47.20 | 532.85±41.98 | 521.61±41.75 | 4.15 | 0.017 |
|  | B (ROP spontaneous regression group) | 542.10±40.19 | 523.80±38.41 | 515.61±32.26 | 6.84 | 0.001 |
|  | C (Non ROP ) | 569.31±44.61 | 519.50±41.53 | 522.35±26.35 | 122.24 | <0.01 |
| ASD/mm | A (ROP post-injection group) | 6.03±0.26 | 6.43±0.24 | 6.59±0.21 | 80.91 | <0.01 |
|  | B (ROP spontaneous regression group) | 6.02±0.23 | 6.45±0.27 | 6.61±0.29 | 83.48 | <0.01 |
|  | C (Non ROP ) | 5.89±0.24 | 6.37±0.30 | 6.59±0.25 | 122.24 | <0.01 |
| LC/mm | A (ROP post-injection group) | 4.04±0.23 | 4.47±0.22 | 4.64±0.19 | 107.85 | <0.01 |
|  | B (ROP spontaneous regression group) | 4.04±0.20 | 4.46±0.24 | 4.60±0.28 | 93.61 | <0.01 |
|  | C (Non ROP ) | 3.95±0.18 | 4.30±0.27 | 4.45±0.20 | 97.12 | <0.01 |
| LT/ACD | A (ROP post-injection group) | 1.94 | 1.6 | 1.47 | 77.8 | <0.01 |
|  | B (ROP spontaneous regression group) | 1.94 | 1.64 | 1.58 | 42.83 | <0.01 |
|  | C (Non ROP ) | 1.96 | 1.89 | 1.88 | 2.75 | 0.066 |

Data are expressed as the mean ±standard deviation.

Eye parameters (ACD, LT, VL, AL, CCT, ASD, LC) were positively correlated with PMA.There was a negative correlation between ACD, ASD, LC and GA (ACD:R=-0.20,P<0.01, ASD:R=-0.15,P<0.01). There was a negative correlation between ACD and BW, and a positive correlation between LT and BW(Table 6). Comprehensive tables 2-6 are available that The development of anterior segment of premature infants in ROP group(group A and group B) was more limited than that in non-ROP groupgroup C. BW, GA and PMA are the main factors affecting the development of ACD, LT, ASD and LC in premature infants with ROP.

**Table 6.** Ocular parameters and their correlation with GA, BW, PMA

| Ocular parameters | GA |  | BW |  | PMA |  |
| --- | --- | --- | --- | --- | --- | --- |
| ACD | r=-0.20 | **p<0.01** | r=-0.10 | **p=0.02** | r=0.36 | **p<0.01** |
| LT | r=0.03 | p=0.49 | r=0.12 | **p=0.006** | r=-0.20 | **p<0.01** |
| VL | r=-0.04 | p=0.32 | r=0.08 | p=0.05 | r=0.15 | **p<0.01** |
| AL | r=-0.08 | p=0.08 | r=0.06 | p=0.20 | r=0.19 | **p<0.01** |
| CCT | r=0.05 | p=0.19 | r=0.04 | p=0.36 | r=-0.13 | **p=0.003** |
| ASD | r=-0.15 | **p<0.01** | r=-0.02 | p=0.62 | r=0.20 | **p<0.01** |
| LC | r=-0.18 | **p<0.01** | r=-0.06 | p=0.15 | r=0.29 | **p<0.01** |

Data are expressed as the mean ± standard deviation.

There was no statistically significant difference in all ocular parameters between the ROP post-injection group and the ROP spontaneous regression group. There were statistically significant differences in ACD, LT, LC and LT/ACD parameters between the ROP post-injection group and the non-ROP group, and between the ROP spontaneous regression group and the non-ROP group.(.(Table1-5))

With age progression, the three groups showed increasing AL, VL, and ACD while CCT decreased across all time points, with no significant differences observed between groups. However, significant variations in LT were detected: the ROP group maintained stable or decreased values, whereas the non-ROP group exhibited marked increases. LT/ACD, which reflects the balance between lens growth and anterior chamber expansion, demonstrated contrasting trends: LT remained unchanged in both the ROP group (including post-injection group and spontaneous regression cases), while it progressively thickened in the non ROP group.Notably, the LT/ACD ratio decreased in ROP cases but remained stable in non ROP cases.(Table 5)

**Figure 1.**
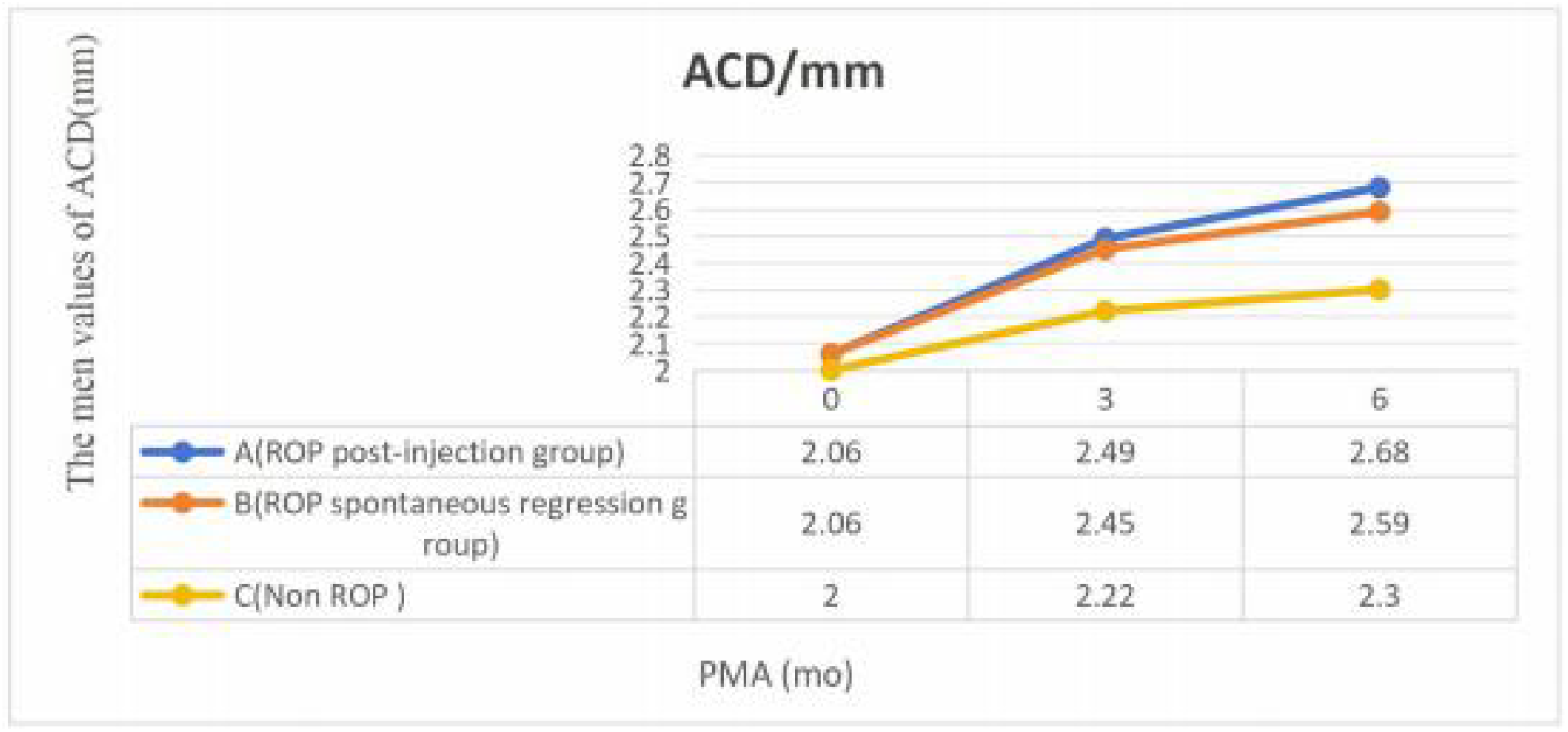
Developmental trend of ACD in three groups of premature infants

**Figure 2.**
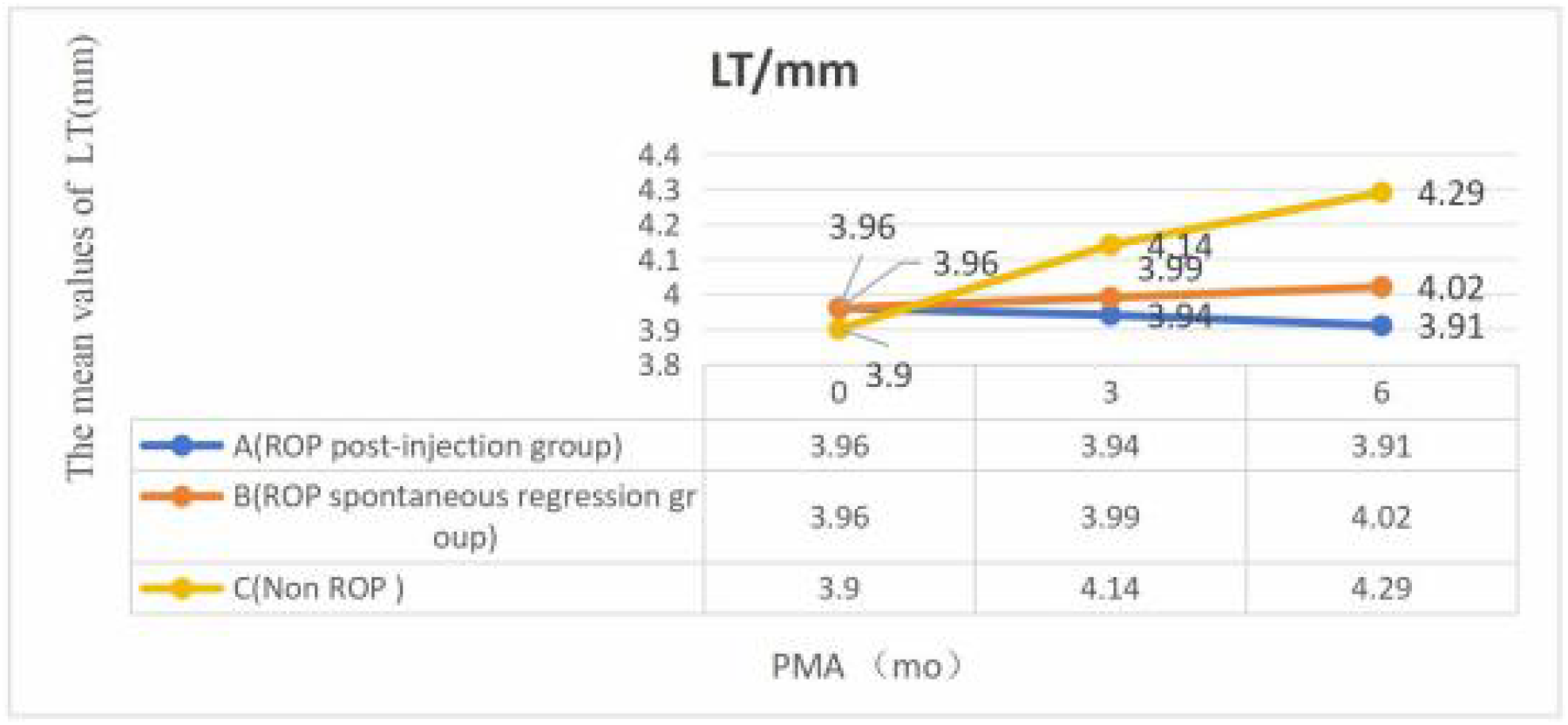
Developmental trend of LT in three groups of premature infants

**Figure3.**
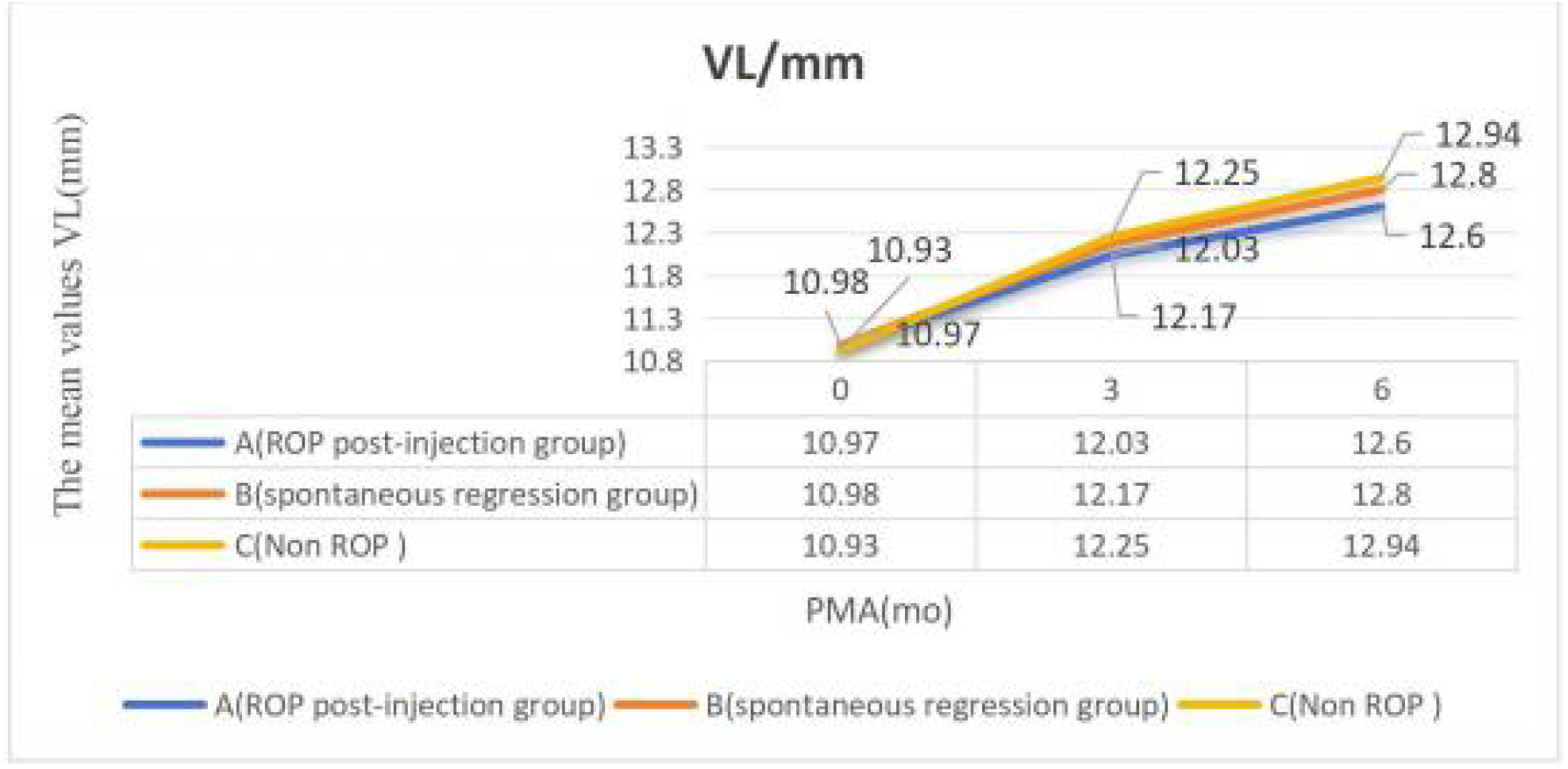
Developmental trend of VL in three groups of premature infants

**Figure4.**
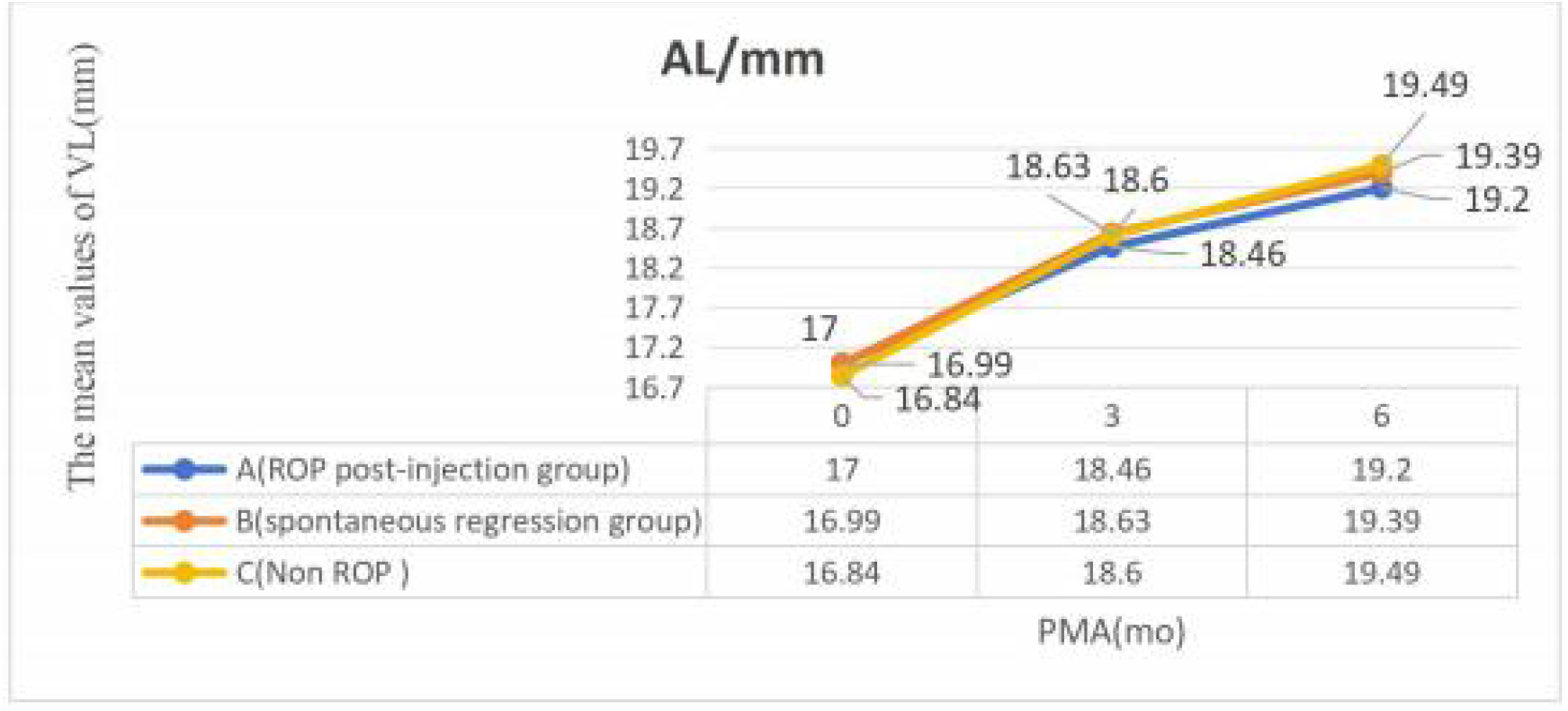
Developmental trend of AL in three groups of premature infants

**Figure5.**
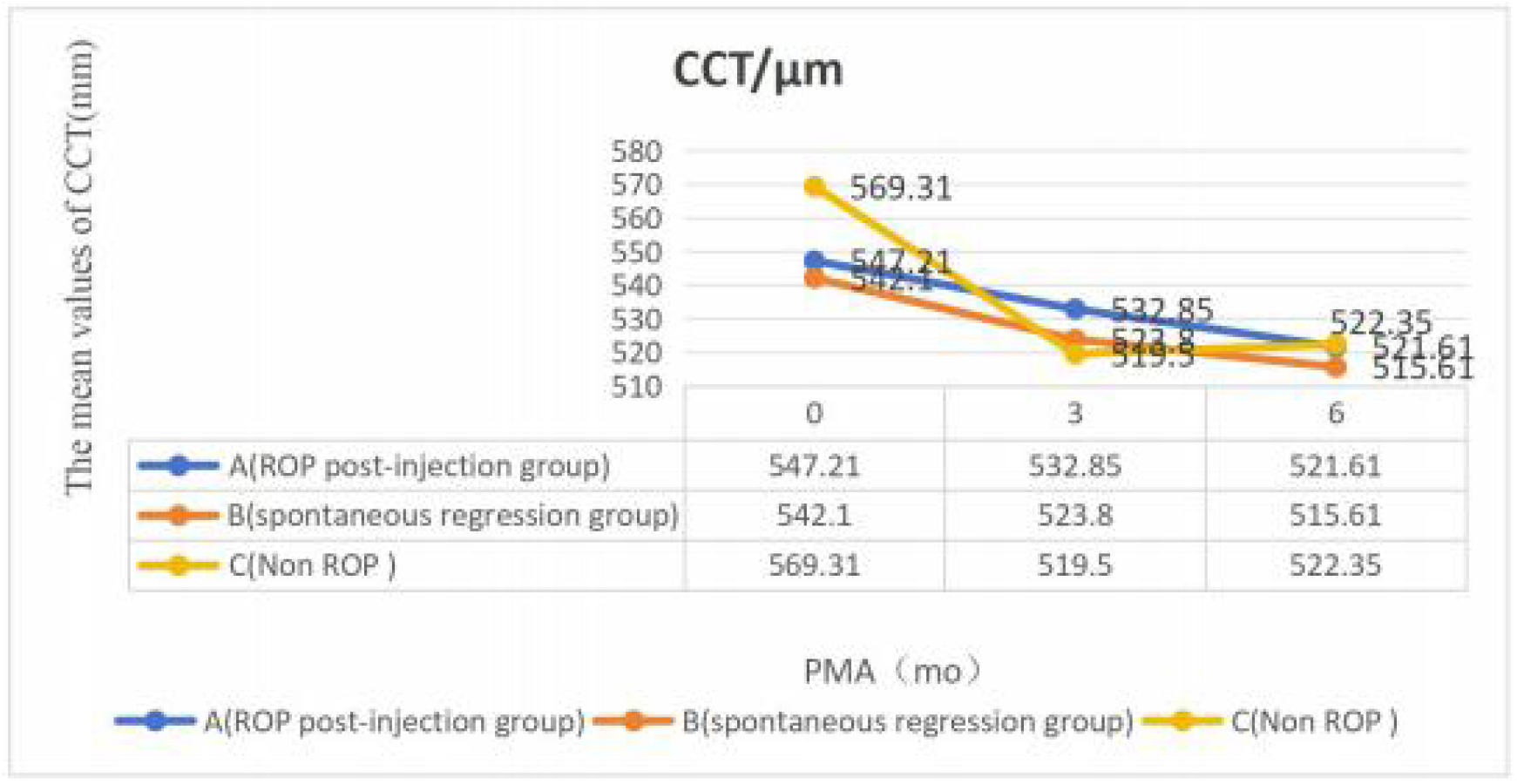
Developmental trend of CCT in three groups of premature infants

**Figure6.**
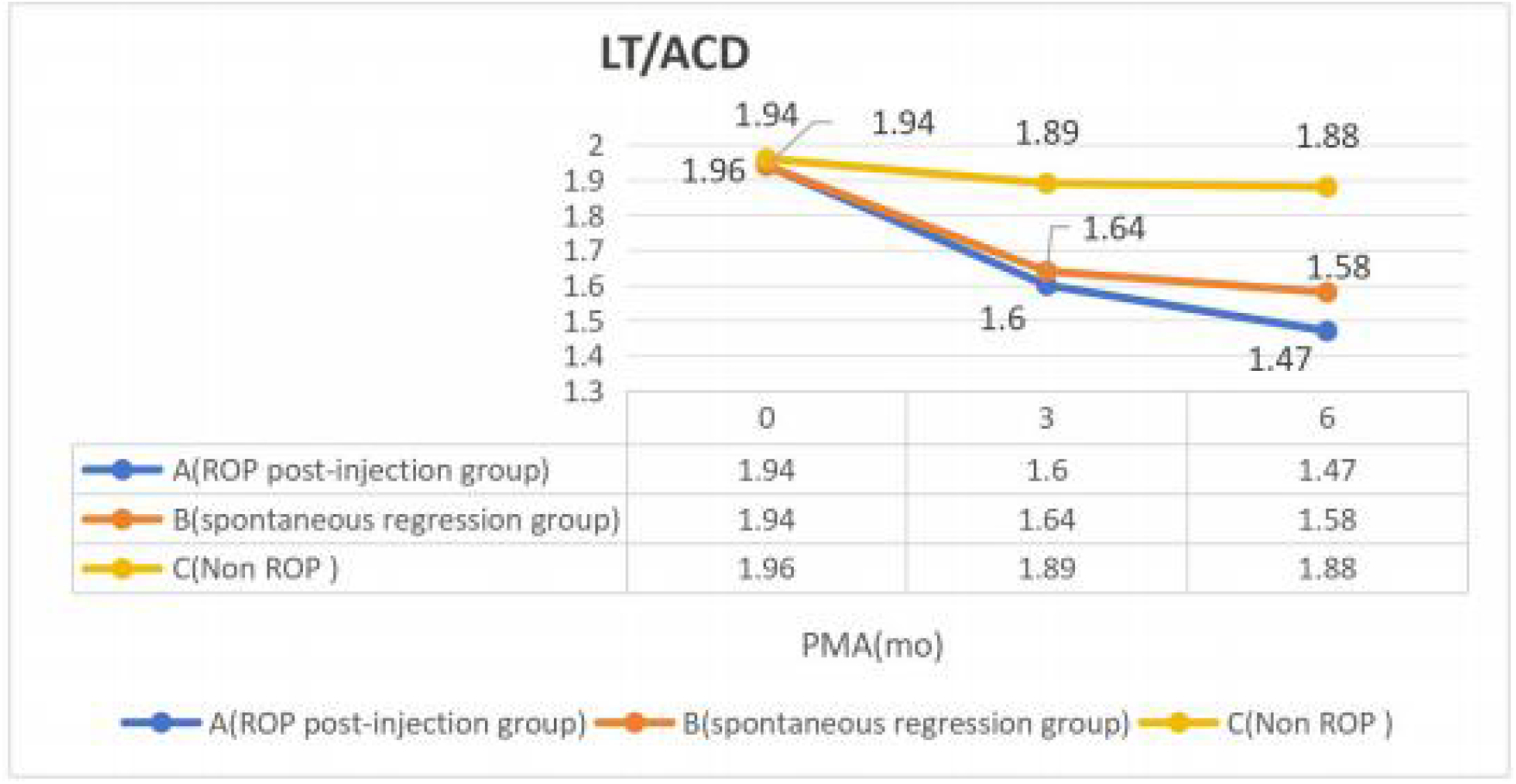
Developmental trend of LT/ACD in three groups of premature infants

**Figure7.**
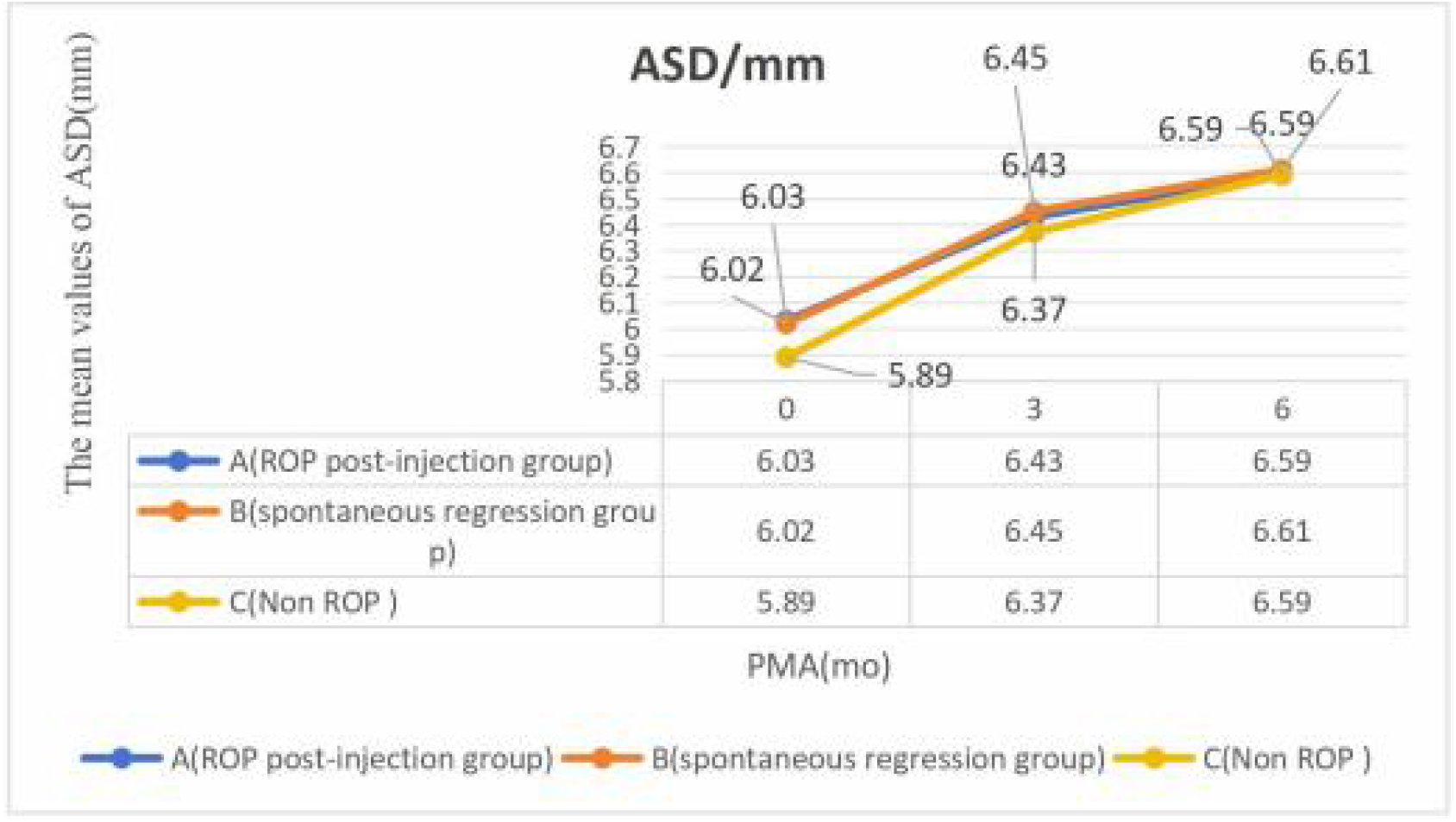
Developmental trend of ASD in three groups of premature infants

**Figure8.**
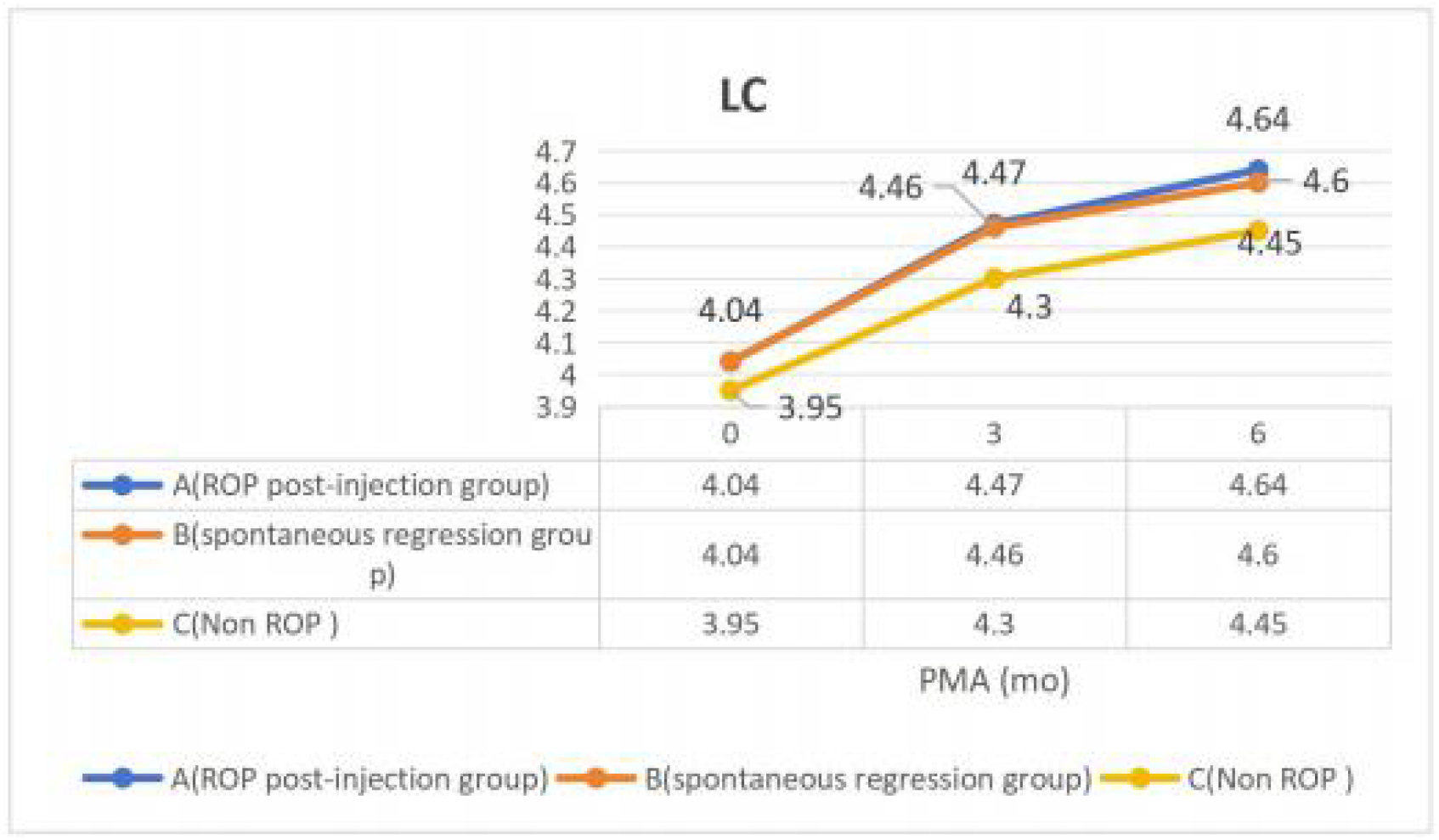
Developmental trend of LC in three groups of premature infants

## 3. Discuss

Jingyun Wang studies suggest that retinopathy of prematurity (ROP) may change and affect the retinal signals needed for eye development and / or sclera reception and response to these signals, delay or interfere with normal scleral development, leading to abnormal development of the anterior segment structure [13]. Zha Y et al believe that the lens of premature infants is relatively spherical near the cornea, and the anterior chamber is shallower and the lens is thicker than that of full-term infants [14]. The lens changes rapidly from birth to the corrected age of 6 months, and the shallower anterior chamber is matched with the increase of lens thickness. Enrique Garcia-Valenzuela et al., believe that the increase of lens thickness leads to the anterior displacement of the anterior surface of the lens so that the depth of the anterior chamber is shallower than that of the full-term infant, and the deepening of the anterior chamber is also related to the thinning of the lens [15]. Yu S J et al., believe that in the development of the human eye, different components of the optical system are developed to prevent ametropia, which is a trend known as “emmetropization” [16].

In this study, the increase of ACD in premature infants with ROP matched with the decrease of LT, and the relationship between LT and ACD was consistent with Enrique Garcia-Valenzuela, which suggested that the shallow anterior chamber matched with the increase of lens thickness, and the decrease of anterior chamber depth was consistent with the anterior displacement of lens surface caused by the increase of lens thickness [17]. Lee et al believe that there is a phenomenon of “one growth and another decline” between LT and ACD, that is, the thicker the LT, the shallower the ACD [18]. Kanclerz and other studies believe that when LT thickens, ACD will decrease, which is consistent with the changes of ROP premature infants in our study [19]. However, the thickening of ACD and LT in premature infants without ROP was consistent with the conclusions of other scholars. Ozdemir study of 361 premature infants found that ACD and LT increased linearly with the post modification age of premature infants [20].

The older the age, the bigger the ASD. In this study, there were significant differences in ASD among the three age points of premature infants in this study. There was significant difference in ASD among the three groups of premature infants in the same age group at PMA 0 month, but there was no significant difference in PMA 3 months and PMA 6 months, and there was no significant difference in VL and AL in all age groups. However, there were significant differences in LC among the three groups of children in each age group. (PMA 0 month F=8. 11,P<0.01 ; PMA 3 month F=8.13,P<0.01,PMA 6 month F=6.36,P=0.003). The emmetropization process of premature infants with ROP is not the same as that of premature infants without ROP. Praveen believes that there is a positive correlation between LT and diopter, that is, the higher the degree of myopia, the thinner LT. In our study, at PMA 0 month,LT:group A=group B>group C, at PMA 3 month and 6 month,LT:group A<group B< group C.The LT of group B and group C had almost no significant change(P<0.05), and the LT of group C increased with age(P > 0.05). Warrier et al think that the lens is thicker in hyperopic people. There is no unified conclusion on the causes of LT inconsistency under different diopters [21]. However, Jonas et al speculated that the reason why LT is thicker in hypermetropia than emmetropia and myopia may be that hyperopia patients need more refractive power when looking near, which induces the thickening of LT [22]. Atchison DA believes that this passive coordination may be the result of emmetropization regulation of the eyeball itself. The reason for this change may be that the fibers in the equatorial part of the lens continue to proliferate, and the newly formed lens fibers are wrapped around the original lens fibers, thus increasing in thickness [23]. Shufelt C believes that when the eye axis has not reached a steady state, LT is mainly affected by the emmetropization regulation of the eyeball itself [24].

In this study, LT was the thickest in group A, the middle in group B and the thinnest in group C in PMA 0 month, while the thickest in group C, the middle in group B and the thinnest in group A in PMA 3 month and 6 month.We speculate that the anterior segment development of premature infants with ROP is blocked in the process of emmetropization compared with those without ROP, and the older the post modification age of LT in group C and group B is, the thicker the LT is, but in group A, the thinner LT is, which may be the result of emmetropization regulation. In this study, there was no significant difference in VL and AL among the three groups of premature infants.. This is consistent with lee et al. [18] and chen [25]. Yebra-Pimentel study suggests that the imbalance between ocular parameters is directly related to the occurrence and development of ametropia. That is, the growth of eyes is a harmonious process in the direction of equator and post-equator, which ensures that the balanced increase of anterior chamber and vitreous body reaches a certain level. Ensure a balance between the anterior and posterior segments. If this level is exceeded, the balance will be lost and ametropia will occur [26]. The key to emmetropization is the negative correlation between the length of eye axis and lens and refractive ability. With the increase of the eye axis, the refractive state of cornea and lens changed. With the deepening of anterior chamber, the curvature of cornea and the thickness of lens began to decrease, and the cornea flattened gradually [27,28].

Cooketal believe that in premature infants, the lens thickens in the first two months after birth and then begins to become thinner [29]. Muttietal et al., believe that the lens of full-term infants begins to thicken from 3 to 9 months [30]. Sivak et al., the study of ametropia in animals has concluded that the lens is passive in the development of ametropia [31]. This decrease in the refractive index of the lens can protect children from long-term myopia during axial tension. The lens becomes thinner due to banded traction caused by the growth of the anterior eyes. As the lens becomes thinner, its curvature flattens [31].

The LC of the three groups of premature infants in our study was 4.04±0.23mm and 4.04±0.20mm VS 3.95±0. 18mm in PMA 0 month, 4.47±0.22 mm and 4.46±0.24mm VS 4.30±0.27 mm in PMA 3 month and 4.64±0. 19 mm and 4.60 ±0.28mm VS 4.45±0.20 mm in PMA 6 month.In the study of Enrique Garcia-Valenzuela, there was no significant difference between ROP injection group and ROP spontaneous regression group[15].In this study, there were significant differences in ROP Post-injection group (group A), ROP spontaneous regression group (group B) and non-ROP group (group C). There were significant differences in LT/ACD among the three age groups of preterm infants after ROP Post-injection (group A) and ROP spontaneous regression group (group B)(P<0.05), but there was no significant difference in non-ROP group (group C)(P<0.05). Fiess A believes that ROP eyes are characterized by growth retardation and abnormality. The abnormal peripheral ROP retina without blood vessels may be the cause of abnormal growth of ROP eyes [28]. Studies on ape eyes have shown that the peripheral retina is the most important for emmetropization [32]. The abnormality (even absence) of the periretinal vascular system in patients with ROP may be due to the retardation of eyeball growth caused by the biological stress of retinopathy. According to Chen TC, ROP lesions are located in the part of the largest eye growth in late fetus and early newborn, and may have a mechanical effect on the anterior sclera and anterior segment [33].

In this study, eye parameters (ACD, LT, VL, AL, CCT, ASD, LC) were positively correlated with PMA (P<0.05). ASD, LC and GA were negatively correlated(P<0.05) (ACD:R=-0.20 P<0.01 ASD:R=-0.15 P<0.01) .ACD was negatively correlated with birth weight(R=-0. 10 P=0.02). There was a positive correlation between LT and birth weight(R=0. 12 P=0.001), which was consistent with Xihui Zhu, MD and other studies. Children with lower body weight tend to have lighter ACD and larger LT [31]. There are many hypotheses about why premature babies are more likely to suffer from myopia than full-term babies. One of the theories [32] is the neuroectoderm hypothesis, especially in infants with ROP, the loss of anterior segmental block will lead to abnormal lens development, resulting in changes in the depth of the anterior chamber due to changes in the development of neuroectoderm in ROP.

In short, there was no difference in VL,AL and ASD in ROP post-injection group (group A), ROP spontaneous regression group (group B) and non ROP group (group C), but there were differences in ACD, LT and LC. We think that ROP possible affects the development of anterior segment of preterm infants, and LT growth retardation of ROP preterm infants may be the main reason for the limitation of anterior segment development.

This study also has some limitations, which is based on the single-center study of the hospital, and the later research should carry out multi-center research.Longer studies and later studies need to be added to the correlation between refractive status and ocular parameters.

## Data Availability

Data is provided within the manuscript or supplementary information files Contacts:Pan pan Ma Telephone:18700084459

## Abbreviations

ACD: Anterior Chamber Depth
AL: Axial Length
LT: Lens Thickness
VL: Vitreous Volume
CCT: Central Corneal Thickness
ASD: Anterior Segment Depth
ROP: Retinopathy of Prematurity
PMA: `Postmenstrual Age` (PMA)
LC: Lens Central
BW: Birth Weight
GA: Gestational Age

## Declarations

### Ethics approval and consent to participate

This study followed the Declaration of Helsinki and was approved by the Ethics Committee of the Northwest Women and Children’s Hospital, and all guardians of the children patients signed informed consent forms.

### Consent for publication

Not Applicable

### Availability of data and materials

Data is provided within the manuscript or supplementary information files Contacts:Pan pan Ma

Telephone:18700084459

### Authors’ contributions

Panpan Ma.Qiong Wu.Xin Wei.Le Zhang Responsible for the writing of the article; The tables and figures

## Acknowledgements

This article is not applicable.

